# Clinico-pathological features in fatal Covid-19 Infection: A Preliminary Experience of a Tertiary Care Centre in North India using Post-Mortem Minimally Invasive Tissue Biopsies

**DOI:** 10.1101/2020.11.12.20229658

**Authors:** Animesh Ray, Deepali Jain, Shubham Agarwal, Shekhar Swaroop, Ayush Goel, Prasenjit Das, Sudheer Kumar Arava, Asit Ranjan Mridha, Aruna Nambirajan, Geetika Singh, S. Arulselvi, Purva Mathur, Sanchit Kumar, Shubham Sahni, Jagbir Nehra, Nazneen, Mouna BM, Neha Rastogi, Sandeep Mahato, Chaavi Gupta, S Bharadhan, Gaurav Dhital, Pawan Goel, Praful Pandey, Santosh KN, Shitij Chaudhary, Vishakh C Keri, Vishal Singh Chauhan, Niranjan Mahishi, Anand Shahi, Ragu R, Baidnath K Gupta, Richa Aggarwal, Kapil Dev Soni, Neeraj Nischal, Manish Soneja, Sanjeev Lalwani, Chitra Sarkar, Randeep Guleria, Naveet Wig, Anjan Trikha

## Abstract

**Background:** The Covid-19 pandemic began in China in December 2019. India is the second most affected country, as of November 2020 with more than 8.5million cases. Covid-19 infection primarily involves the lung with severity of illness varying from influenza-like illness to acute respiratory distress syndrome. Other organs have also found to be variably affected. Studies evaluating the histopathological changes of Covid-19 are critical in providing a better understanding of the disease pathophysiology and guiding treatment. Minimally invasive biopsy techniques (MITS/B) provide an easy and suitable alternative to complete autopsies. In this prospective single center study we present the histopathological examination of 37 patients who died with complications of Covid-19.

**Methods:** This was an observational study conducted in the Intensive Care Unit of JPN Trauma Centre AIIMS. A total of 37 patients who died of Covid-19 were enrolled in the study. Post-mortem percutaneous biopsies were taken by the help of surface landmarking/ultrasonography guidance from lung, heart, liver, and kidneys; after obtaining ethical consent. The biopsy samples were then stained with haematoxylin and eosin stain. Immunohistochemistry (IHC) was performed using CD61 and CD163 in all lung cores. SARS-CoV-2 virus was detected using IHC with primary antibodies in selected samples. Details regarding demographics, clinical parameters, hospital course, treatment details, and laboratory investigations were also collected for clinical correlation.

**Results:** A total of 37 patients underwent post-mortem minimally invasive tissue sampling. Mean age of the patients was 48.7years and 59.5% of them were males. Respiratory failure was the most common complication seen in 97.3%. Lung histopathology showed acute lung injury and diffuse alveolar damage in 78% patients. Associated bronchopneumonia was seen in 37.5% patients and scattered microthrombi were visualised in 21% patients. Immunostaining with CD61 and CD163 highlighted megakaryocytes, and increased macrophages in all samples. Immunopositivity for SARS-CoV-2 was observed in Type II pneumocytes. Acute tubular injury with epithelial vacuolization was seen in 46% of the renal biopsies but none of them showed evidence of microvascular thrombosis. 71% of the liver tissue cores showed evidence of Kupfer cell hyperplasia. 27.5% had evidence of submassive hepatic necrosis and 14% had features of acute on chronic liver failure. All the heart biopsies showed non-specific features such as hypertrophy with nucleomegaly with no evidence of myocardial necrosis in any of the samples.

**Conclusions:** The most common finding in this cohort is the diffuse alveolar damage with demonstration of SARS-CoV-2 protein in the acute phase of DAD. Microvascular thrombi were rarely identified in the lung, liver and kidney. Substantial hepatocyte necrosis, hepatocyte degeneration, Kupffer cell hypertrophy, micro, and macrovesicular steatosis unrelated to microvascular thrombi suggests that liver might be a primary target of Covid-19. This study highlights the importance of MITS/B in better understanding the pathological changes associated with Covid-19.

## Introduction

The beginning of 2020 witnessed the expansion of the COVID-19 pandemic, which began in the Wuhan district of China, in December 2019. In India, the first case was documented on 30^th^ January 2020. As of 8^th^ November 2020, the epidemic in India continues to accelerate with 85,41,173 cases and 1,26,525 deaths reported.Severe acute respiratory syndrome coronavirus 2 (SARS-Cov-2) is a positive sense, enveloped, single-stranded RNA virus belonging to the Coronavirus family.(1)

The spectrum of COVID-19 infection ranges from asymptomatic disease, mild influenza-like illness, to severe pneumonia, acute respiratory distress syndrome (ARDS), multiorgan failure, and death. Fever is the most common presenting symptom (87.3%) followed by cough (58%) and shortness of breath (38.3%). Radiographic findings include bilateral pneumonia and ground-glass opacities. Roughly 28% of the patients go on to develop acute respiratory distress syndrome and the case fatality rate is around 7%.(2) Other organs have also been found to be variably affected like heart, kidneys, and liver with patients developing complications like arrhythmias, acute kidney injury, and coagulopathy although the pathogenesis is still unclear.

Studies that evaluate histopathological changes associated with SARS CoV-2 are critical towards a better understanding of the pathophysiology of this disease. Minimally invasive post-mortem tissue sampling or biopsies (MITS/B) provide an effective, acceptable, and technically less challenging alternative to complete autopsies in fatal COVID-19 patients, with less chance of transmission of infection. Studies done so far show predominant findings of diffuse alveolar damage in the lung accompanied by capillary congestion and microthrombi.(3)

In this prospective single-center study, we present the histopathological findings of various organs of 37 patients, who died due to complications of COVID-19 in order to provide insights into the disease pathogenesis, pathological changes in various organ systems and suggestions for treatment.

## Methodology

### Study design and patients

This was an observational study, conducted in the ICUs of Jai Prakash Narayan Apex Trauma Center (JPNATC), AIIMS, New Delhi between May to August 2020.

A total of 37 patients who died of Covid-19 were enrolled in the study. All of them were diagnosed antemortem by RT-PCR/TrueNAT/Antigen testing. Ethical consent for post-mortem tissue sampling was obtained from the next of kin as per institutional guidelines. The study was approved by the Institute Ethics Committee (Ref No: IEC-536/05.06.2020).

Demographic data including age, sex, residence, occupation, contact history were recorded. Relevant clinical data including presenting complaints, physical examination, hospital course, laboratory and radiological investigations, treatment given along with the probable cause of death were recorded in an online proforma.

### Biopsy technique

Post-mortem minimally invasive tissue sampling was done within 2 hours of death (cessation of circulation and electrocardiogram showing asystole). Sampling was attempted from lungs, heart, liver, spleen, and kidneys (all organs when feasible, otherwise most organs were targetted). Percutaneous biopsies were done using B-BARD® biopsy gun with or without ultrasonography guidance, using surface land markings. The site for biopsy was cleaned with chlorhexidine, followed by insertion of the charged biopsy gun through the skin into the target organ. The gun was fired thereafter, thus collecting a 1-3cm sized tissue core/s. 4-6 biopsy specimens were collected from each organ as per feasibility by using the same skin orifice. Lung specimens were collected from the mid-axillary approach, heart specimens from the left para-sternal approach, liver/spleen specimens from the lower mid-axillary approach, and kidney specimens from the posterior or lumbar approach.

The tissue samples were then transferred to vials containing 10% buffered formalin(10 X volume of the specimen),which was freshly prepared from 40% formaldehyde after adding 1 part of formalin and 9 parts of water with 4.5g/L monobasic sodium phosphate and 6g/L dibasic sodium phosphate. The samples were transported to the lab on the same day at room temperature while maintaining standard precautions.

### Histopathological processing and examination

The biopsy samples were grossed in Biosafety cabinet-2 (BSC-2). The slides obtained from paraffin-embedded blocks were stained by routine hematoxylin and eosin stain. Morphology of the organ tissue (lung, heart, kidney, liver, brain) submitted was studied and recorded by pathologists having experise in a particular organ system. Special stains (AFB, SM, PAS) were done to supplement the basic microscopic findings when needed. Immunohistochemistry for CD163 (MA5-11458, Invitrogen, 1:200) and CD61 (MA1-80862, Invitrogen, 1:50) was performed in all lung cores, while SARS-CoV-2 virus was detected by immunohistochemistry using primary antibodies directed against SARS nucleoprotein (B46F clone, Invitrogen, Catalog # MA1-7404, 1:400), in selected lung samples.

## Results

MITS/B was performed for 37 patients, after obtaining informed consent as detailed earlier. Baseline clinical data were obtained for all patients. Lung tissue cores were taken for 33 patients, liver tissue cores for 29, kidney tissue cores for 11 patients, and heart tissue cores for 15 patients.

### Clinical characteristics (Table 1)

The mean age of the deceased patients was 48.7 years. 59.5% of them were males. About 95% of them had one or more co-morbidities, most common being hypertension followed by diabetes and chronic kidney disease. 35.1% of the patients presented with severe acute respiratory illness (SARI)and 65.6% of them needed oxygen therapy at presentation. The median duration of hospital stay was 7 days with all patients requiring invasive ventilation during the hospital course. Anemia was seen in 85.3% of patients and lymphopenia in 72% of them. 48.1% of the patients had typical findings of COVID-19 on chest radiograph characterized by peripheral-predominant lowerlobe opacities.

Intravenous (IV) broad-spectrum antibiotics were given in all 37 patients, started empirically on day 1 of admission (as per institutional protocol). IV high dose steroids were used in 25 patients (67.5%), while anticoagulation was given to 16 patients (43.2%). Remdesivir was given to 3 patients and 3 patients received tocilizumab.

All patients had one or more complications, with respiratory failure being most common, seen in 36 cases (97.3%). Secondary infections and sepsis syndrome, based on clinical and laboratory parameters, were seen in 21 patients (56.7%). Other complications included arrhythmias were seen in 4 patients, coagulopathy and clinically significant bleeding in 4 cases, cytokine release syndrome in 8 cases, acute ischemic stroke in 1 patient, and deep vein thrombosis (DVT) in 1 patient.

### Pulmonary histopathology (Figure 1)

Lung parenchyma was sampled in 32 patients (Figure2).

**Figure 1.**
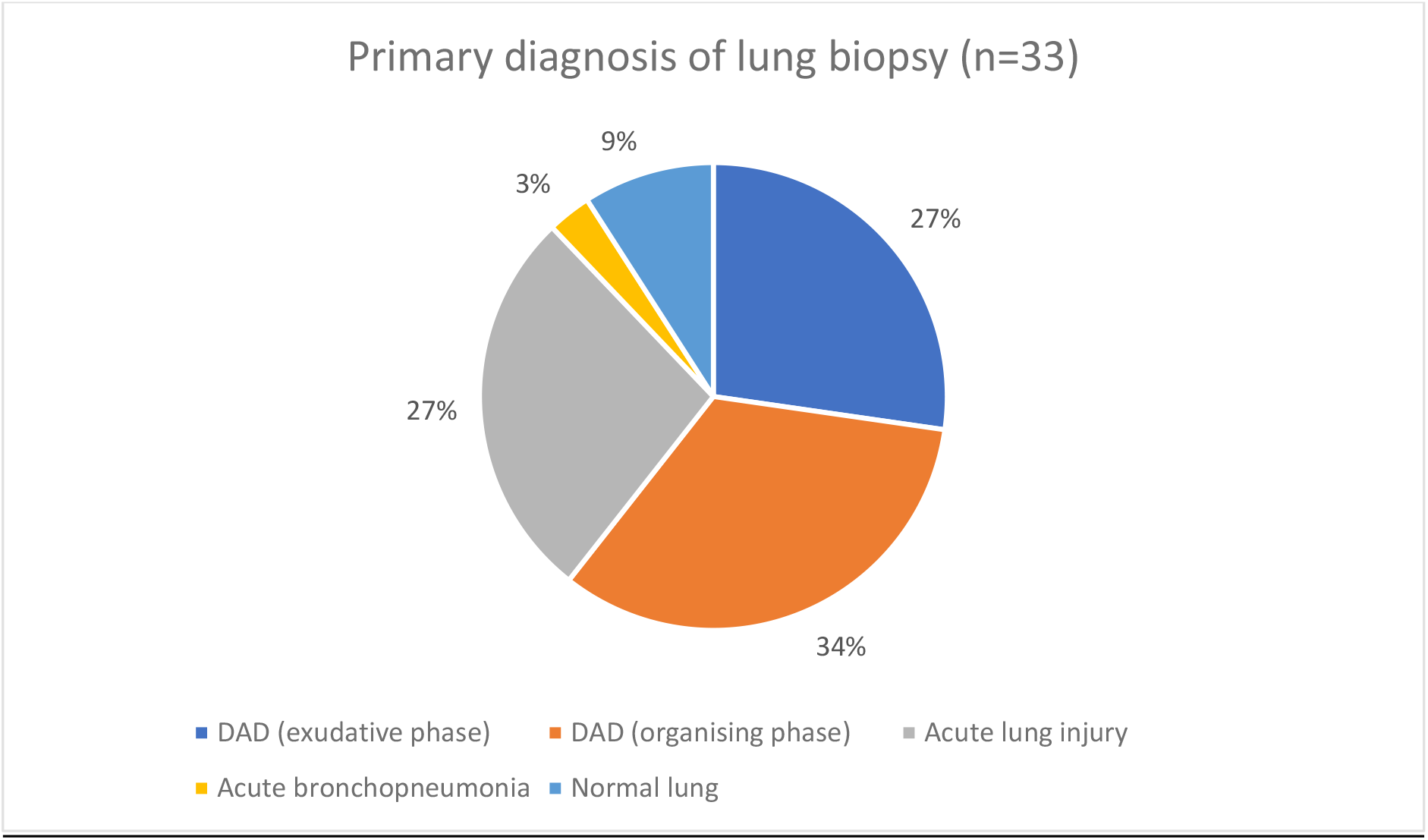

**Figure 2:**
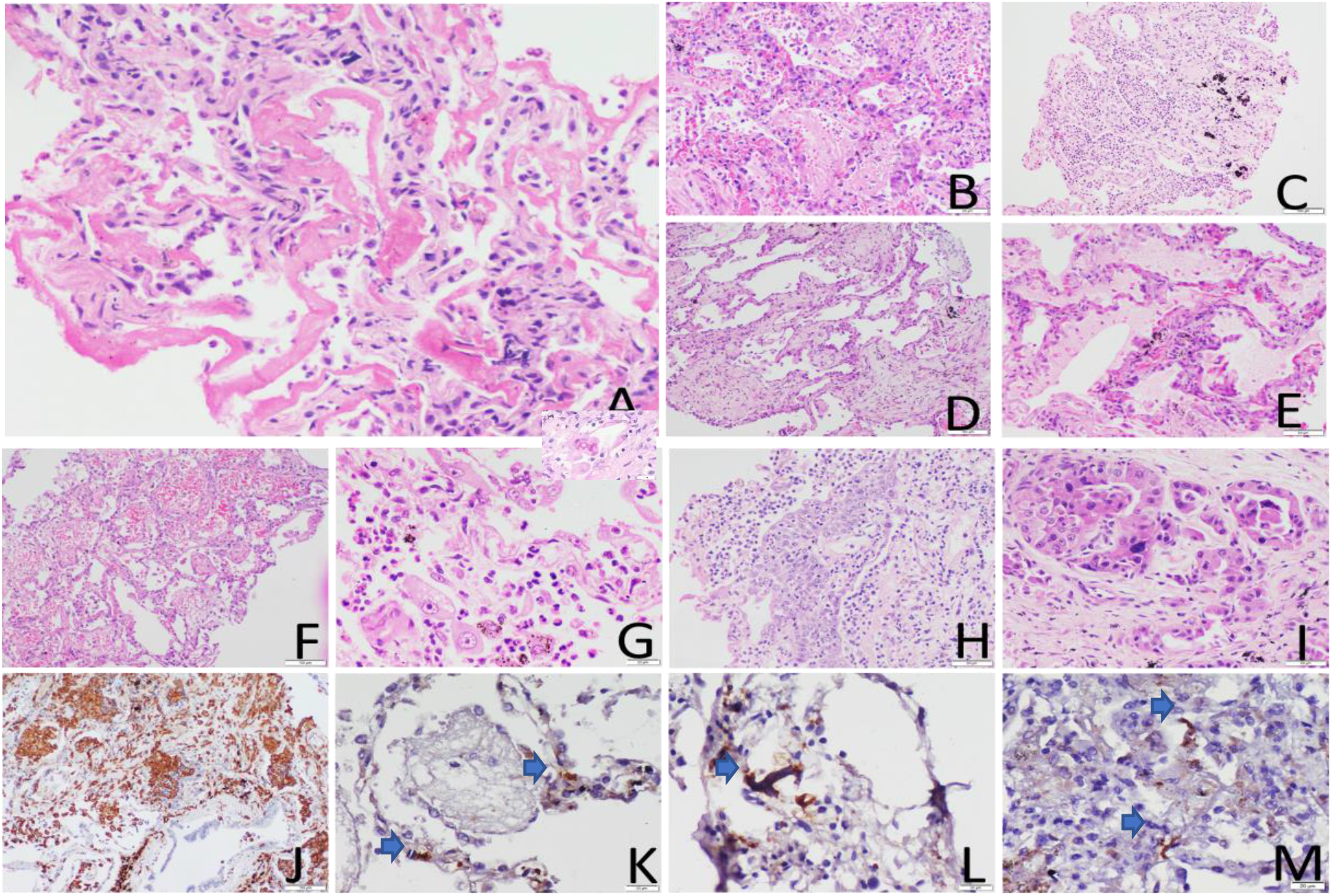
**(A)** Diffuse alveolar damage – exudative phase with hyaline membranes. **(B)** Acute lung injury with engorged capillaries. **(C)** Acute bronchopneumonia. **(D)** Diffuse alveolar damage – organizing phase.**(E)** Pulmonary edema **(F)** Alveolar hemorrhage **(G)** Hyperplastic pneumocytes with inclusion like nucleoli; Inset shows cytoplasmic hyaline in Type 2 pneumocytes **(H)** Pneumocyte hyperplasia**(I)** Adenocarcinoma. **(J)** Anti-CD163 highlights infiltration in acute organizing phase of DAD. (**K)** Anti-CD61 immunostaining highlights occasional microthrombi in alveolar capillaries **(L)**and also megakaryocytes. **(M)**Immunostaining using anti-SARS-CoV-2 antibody is positive in alveolar epithelial cells.

Acute lung injury and diffuse alveolar damage were seen in most of the patients (25/32, 78%) with morphological evidence of only acute lung injury in 4 patients, exudative phase of diffuse alveolar damage (DAD) with hyaline membranes in 10 patients, and organizing phase of DAD with interstitial fibroblastic proliferation and/or intra-alveolar fibroblastic plugs in 11 patients. One biopsy categorized as organizing DAD also showed normal lung parenchyma in some focal areas. Associated bronchopneumonia was seen in 12 of these patients, the majority associated with the organizing phase of DAD (7/12), with occasional cases co-existing with acute exudative DAD (3/12) and acute lung injury (2/12). Bacterial colonies, indicating superimposed bacterial infection, were observed in one of the cases associated with organizing DAD. Other associated findings observed in these patients include incidental carcinoma (n=2), alveolar hemorrhage (n=2), and pulmonary edema (n=1).

In the remaining 7 patients without evidence of acute lung injury or DAD, 3 biopsies showed normal lung parenchyma, 2 showed interstitial chronic inflammatory infiltration, 1 showed tiny lung parenchyma with fibrin deposition, and 1 showed features of acute bronchopneumonia with bacterial colonies.

Scattered microthrombi were seen in 21% (7/32) of the patients, all observed in lung biopsies showing organizing DAD (n=6) or acute DAD (n=1).

Scattered megakaryocytes and hyperplastic pneumocytes were observed in 7 cases and 5 cases respectively. (Table 2).

**Table 1.**
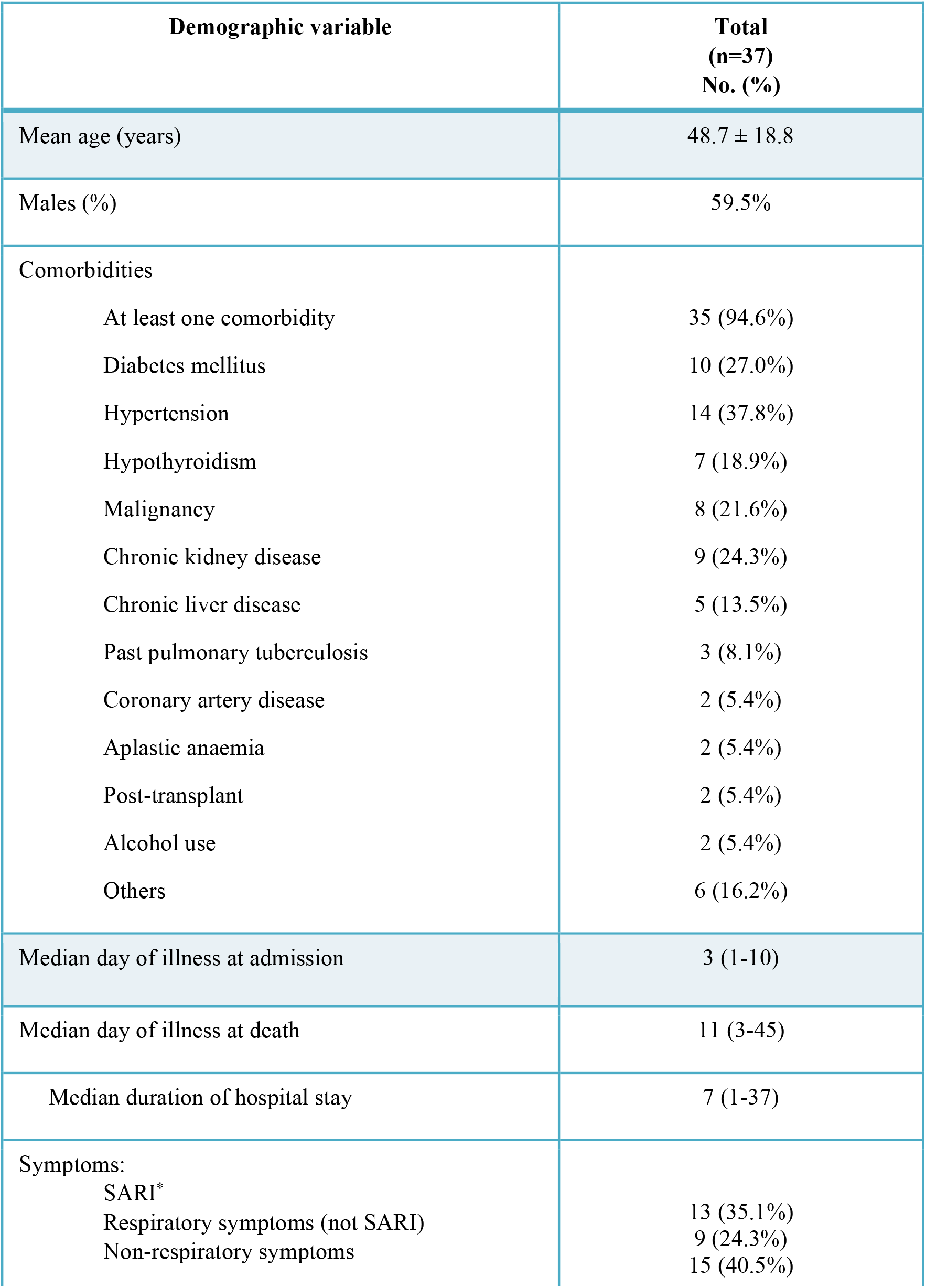
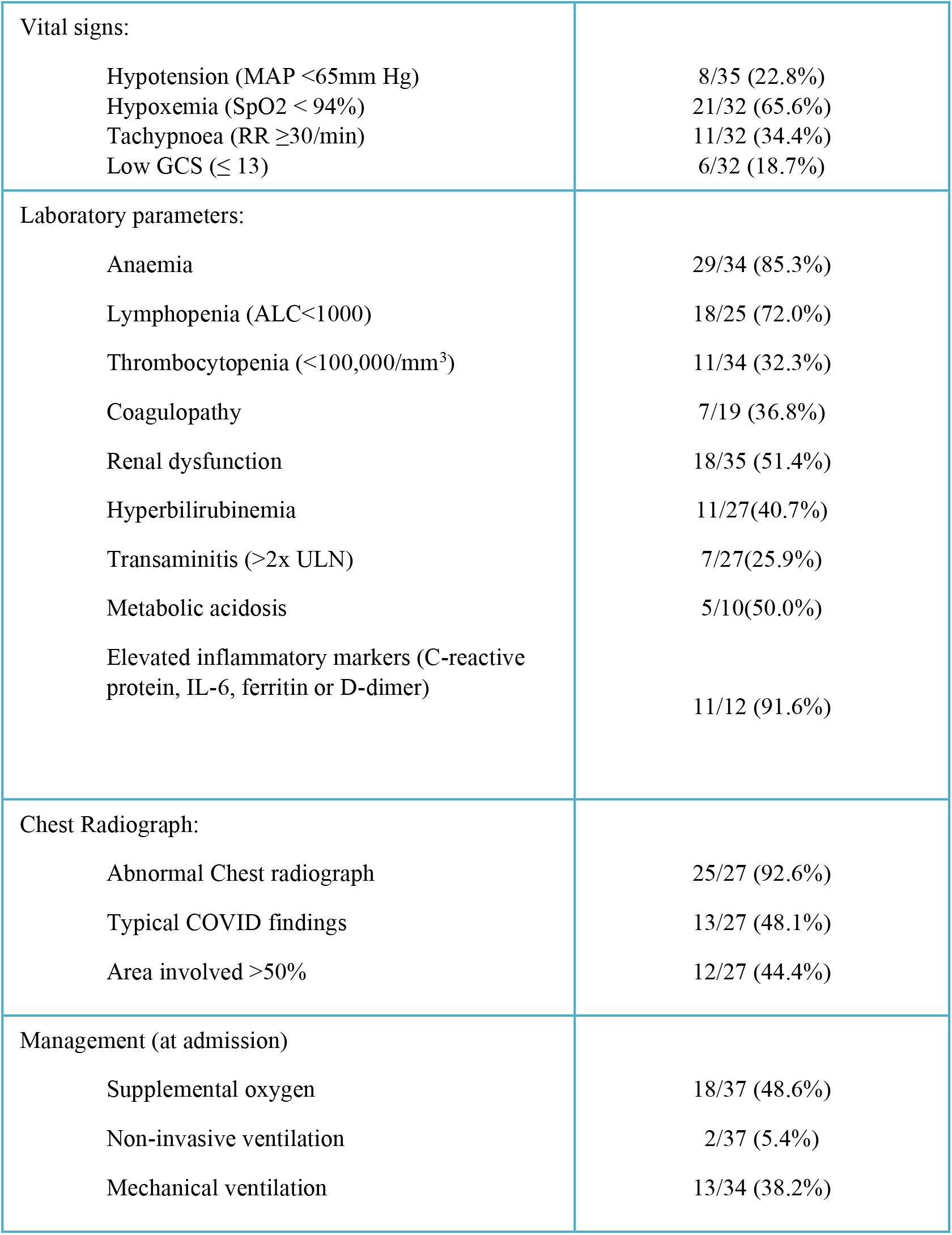

**Table 2.**

|  |  |
| --- | --- |
| <u>Vascular changes in the lung</u> | <u>N=33</u> |
| Microthrombi | 3(9%) |
| Engorged capillaries | 3(9%) |
| Normal | 27(82%) |

**Table 3.**

|  | <u>N=25</u> | <u>Vascular changes in Liver</u> | <u>N=25</u> |
| --- | --- | --- | --- |
| Acute liver failure | 1 | Thrombi/obliteration- Portal vein | 1 |
| Acute on chronic liver failure | 3 | Thrombi/obliteration-Perisinusoidal | 1 |
| Changes of heart failure | 4 | Thrombi/obliteration-Hepatic vein | 1 |
| Submassive hepatic necrosis | 6 | Thrombi/obliteration-HV+PS | 1 |
| Non-cirrhotic portal fibrosis | 3 | Thrombi/obliteration-PV+PS | 1 |
| NAFLD | 1 | Normal | 20 |
| Macrovesicular steatosis | 3 |  |  |
| Microvesicular steatosis | 1 |  |  |
| Normal | 3 |  |  |

Immunostaining for CD163 showed increased macrophages in all samples while CD61 immunostaining highlighted megakaryocytes and microthrombi.

Immunohistochemistry for SARS-CoV-2 nucleoprotein was performed in six biopsies and immunopositivity in Type II pneumocyte was observed in one case of acute DAD and none of the organizing DADs. No immunopositivity was observed in endothelial cells, bronchial epithelial cells, or in the hyaline membranes.

### Renal histopathology (Figure 3)

The most common finding was diffuse acute tubular injury with epithelial vacuolization seen in 46% of the patients (Figure 4). None of the cases demonstrated microvascular thrombosis. Other histological findings noted were of the underlying kidney disease including advanced diabetic glomerulosclerosis (n=1) and early diabetic/hypertensive changes (n=1). Two patients had history of renal transplantation, of which one had a concomitant chronic active T cell and antibody-mediated rejection and the other had focal segmental glomerulosclerosis.

**Figure 3.**
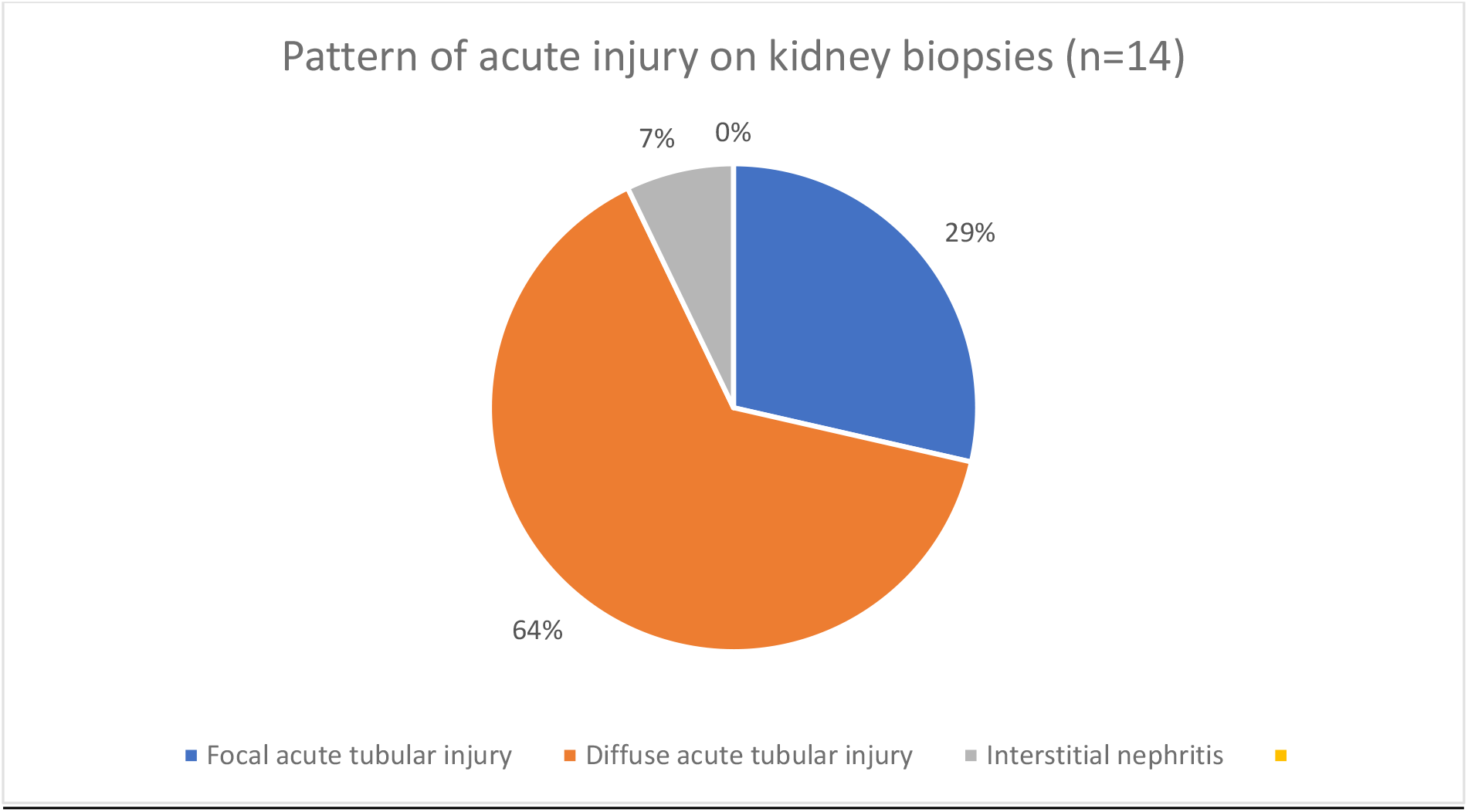

**Figure 4:**
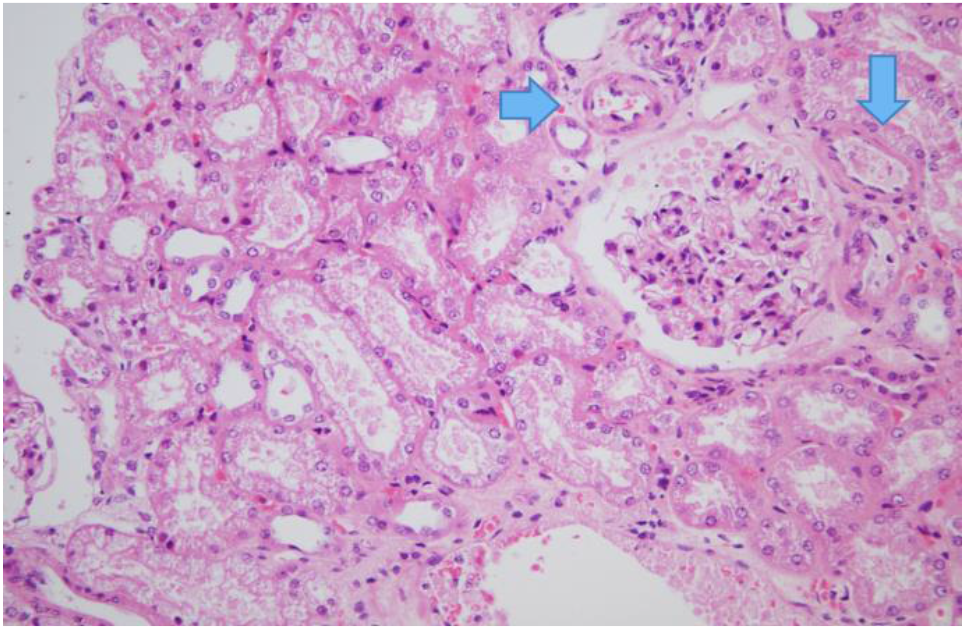
Kidney biopsy showing acute tubular injury.

### Liver histopathology (Table 3)

Liver biopsy cores were obtained from 29 of these 37 patients. In 8 patients (27.58%) acute submassive hepatic necrosis was identified, while in 4 (13.79%) biopsies features of acute on chronic liver failure (ACLF) were identified with the presence of hepatic necrosis (Figure 5). Four of the biopsies showing acute hepatic necrosis also showed prominent microvesicular steatosis. In one of the liver biopsies showing features of ACLF, the histological features were like an acute exacerbation of autoimmune inflammation. However, the most common histological finding was Kupffer cell hypertrophy seen in 21 (72.41%) biopsies. In 3 biopsies (10.3%) there were features of non-cirrhotic portal fibrosis (NCPF), in the form of portal vein thrombosis in 2 biopsies with or without focal sinusoidal microthrombi. Portal inflammation, interface hepatitis, or lobular necroinflammatory activity were not prominent in any of our cases. While significant portal mononuclear cell infiltrates were identified in 13 (44.82%) biopsies, in only 3 biopsies the portal inflammation was dense, while in only 4 of these biopsies the portal inflammation was moderate. Lobular inflammation was identified only in 5 (17.24%) cases (Figure 6). In 4 biopsies (13.79%) there was centrizonal hemorrhagic necrosis, histologically suggestive of heart failure. Macrovesicular steatosis in zone 3 and zone 2 were identified in 4 (13.79%) and diffuse macrovesicular steatosis was noted in one biopsy (Figure 6). Out of the liver biopsies showing macrovesicular steatosis, two of these patients had a history of chronic hypothyroidism, while all of them also got steroids as part of their management regimen during the current hospital course of the COVID-19 related episode.In two of the biopsies from patients with hypothyroidism, we also identified prominent nuclear glycogenization (Figure 6). Liver biopsy cores from 4 of these deceased were within normal histological limits. The biopsies showing hepatic necrosis had a varied pattern. while zone 3 predominant necrosis was seen in 8 biopsies (Figure 6H), diffuse transacinar necrosis was noted in 2 and focal irregular necrosis pattern was identified in 3 of these biopsies. Other features of hepatocyte damage viz. ballooning, acidophil bodies,and Kupffer cell hyperplasia were also prominent in most of the liver biopsies, except the histologically normal biopsies (Figure 6). Intracanalicular and hepatocyte cytoplasmic cholestasis were identified in 7 biopsies (24.13%) and ductular cholestasis was identified in one biopsy with other features of associated sepsis-related changes.

**Figure 5:**
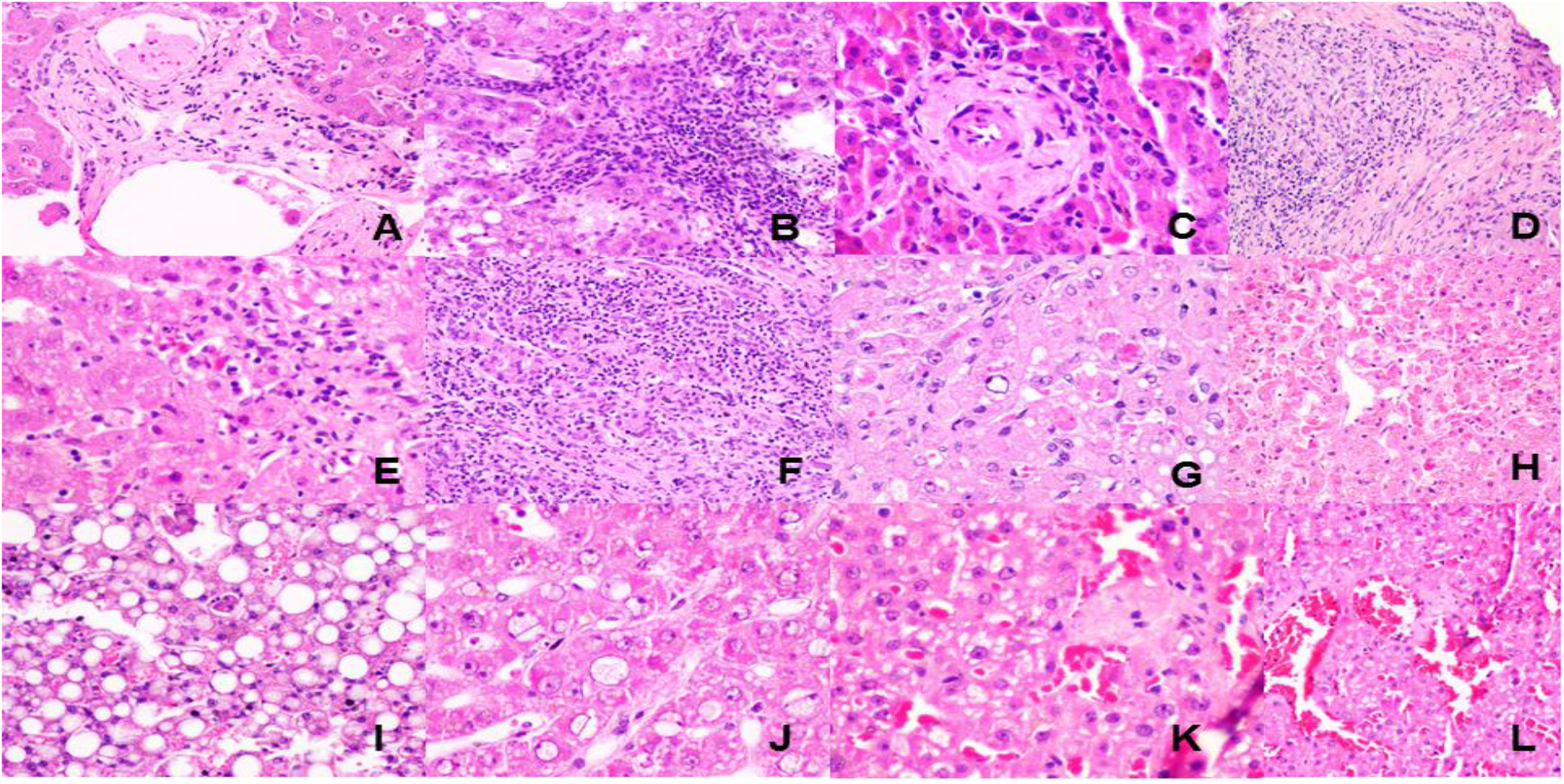
**(A)** Normal portal tract. **(B)** Moderately dense portal mononuclear cell infiltrate with interface hepatitis. **(C)** Portal tract with absence of bile duct. **(D)** Thrombosed and recanalized portal vein. **(E)** Spotty necrosis. **(F)** Lobular lymphoplasmacytic cell infiltrate and diffuse destruction of hepatocytes in a case of acute autoimmune flare. **(G)** Mallory Denk bodies, necrotic hepatocytes, nuclear glycogenization and macrovesicular steatosis in a case of alcoholic cirrhosis. **(H)** Centrizonal hepatocyte necrosis. **(I)** Macrovesicular steatosis and Kupffer cell hypertrophy. **(J)** Microvesicular steatosis. **(K)** Focal sinusoidal fibrosis. **(L)** Thickening and fibrosis of central hepatic vein.

**Figure 6:**
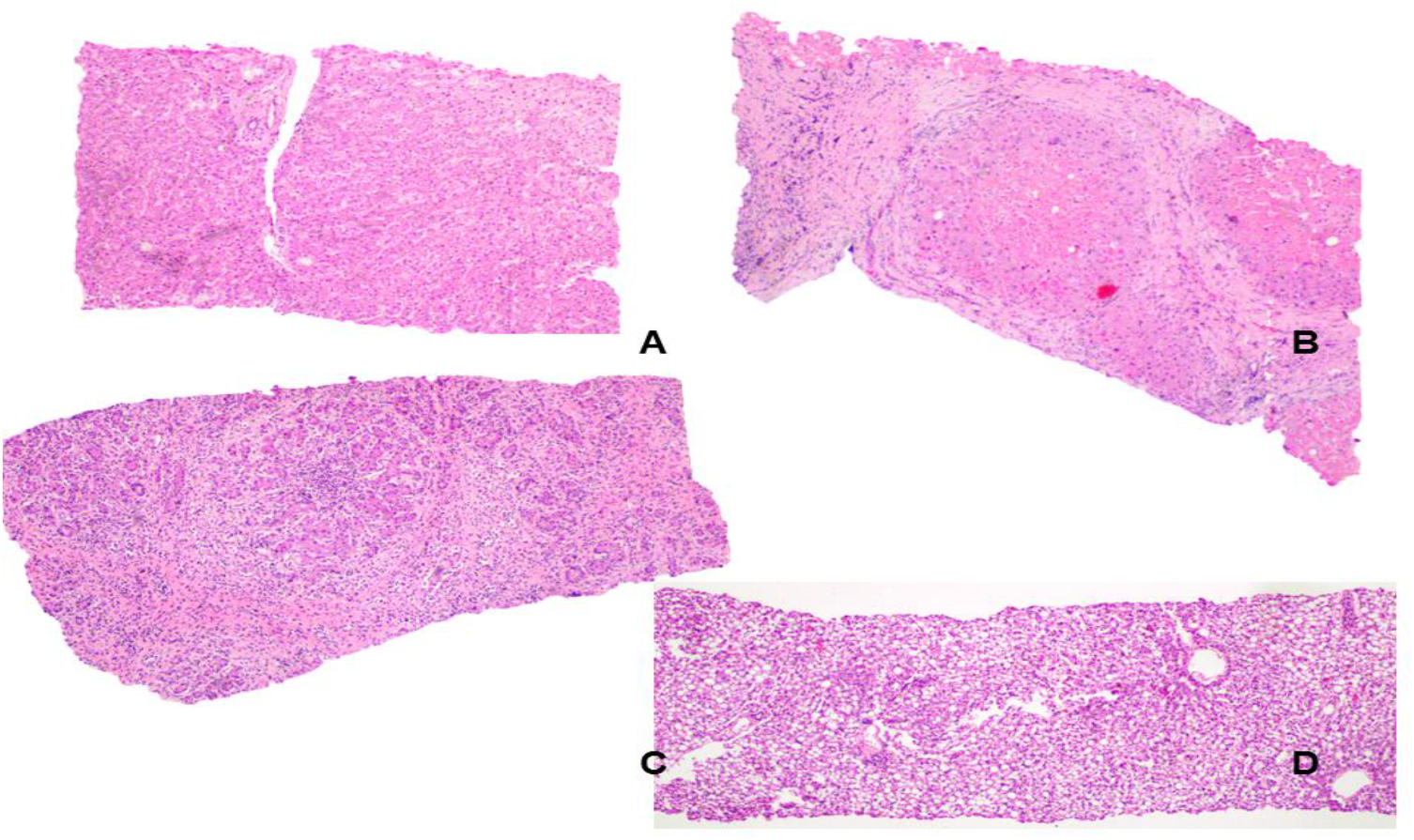
**(A)** Liver with maintained lobular architecture. **(B)** Cirrhotic liver with activity. **(C)** Cirrhosis with activity and severe autoimmune flare. **(D)** Maintained architecture and diffuse macrovesicular steatosis

### Myocardial histopathology

All the heart biopsies showed non specific histopathological features like mild hypertrophy with nucleomegaly of the cardiac myocytes. None of the biopsy samples showed evidence of myocyte damage or necrosis, which is the hall mark of myocarditis. However only one biopsy showed presence of few lymphocytic infiltrtation in the interstitium without myocyte damage indicative of borderline myocarditis.

## Discussion

Minimally invasive autopsies (MIA) or MITS/B are a simplified method of conducting post-mortem sampling, originally devised to investigate the causes of death in low-resource settings. Due to the potential risks associated with conducting traditional post-mortem examinations in COVID-19 patients, this method has been adopted to study morbid pathological changes in COVID-19 patients.(4,5) This method besides being effective(6,7) also has the added advantage of being more acceptable(8) and less time-consuming than elaborate autopsies. In this series of post-mortem samples from 37 patients from a single tertiary care center in India, we studied the morbid pathological changes in COVID-19 patients.

The typical findings described in pulmonary histopathology of COVID-19 patients include epithelial,(3) vascular,(9) fibrotic(10) and other changes. The epithelial changes described include diffuse alveolar damage with or without hyaline membranes, metaplasia of alveolar epithelium, desquamation/ reactive hyperplasia of pneumocytes, viral cytopathic changes, and multinucleated giant cells.(3)Common vascular changes include capillary congestion, thrombosis in microvasculatures, alveolar hemorrhage, capillary changes(proliferation, thickening, fibrin deposition, endothelial detachment), peri or intra-vascular inflammatory infiltrates.(6)The fibrotic changes include interstitial fibrosis or microcystic honeycombing.(3)Other findings that can be encountered include interstitial or intra-alveolar inflammatory infiltrates or edema.(3) In our series, the most common histopathological pattern seen on lung samples was DAD (diffuse alveolar damage), with nearly equal numbers of patients showing DAD in the acute exudative phase and organizing phase. In pathological examination of acute respiratory distress syndrome(ARDS) patients, DAD(exudative) is known to be the predominant feature till the 6^th^ day followed by a transition to organising phase till the end of the second week before progressing to fibrotic stages. (11) The median duration of hospital course before death was 7 days in our series, which explains the predominance of DAD features. Associated bronchopneumonia, particularly in association with organizing DAD, was observed in 13 patients, of which obvious bacterial etiology in the form of bacterial colonies was observed in only two cases. A similar high frequency of bacterial culture-negative bronchopneumonia has been observed previously(12) suggesting that this may be a pathological manifestation of the virus itself. The incidence of vascular changes were only 21%, which was less than that reported.(12,13) Several factors could have contributed to this observation including regular use of anticoagulants in the cohort as a part of national policies(14), lower thrombotic complications in the population studied (due to genetic(15) or climatic factors(16)) as well as sampling errors. We were able to demonstrate SARS-CoV-2 nucleoprotein in one case among 6 cases tested in Type 2 pneumocytes using immunohistochemistry. A recent study has demonstrated specific detection of SARS-CoV-2 in areas of acute DAD and the hyaline membranes while areas of organizing DAD did not show detectable viral antigen indicating viral clearance in the later stages of ARDS (17). In support of this observation, we observed immunostaining for viral antigen in areas of acute DAD.

Histopathological examination of liver in COVID-19 patients had typically revealed mild steatosis, focal hepatic necrosis, Kupffer cell hyperplasia, and sinusoidal dilatation as reported in literature.(4,18,19) Less commonly reported findings include inflammation of portal/sinusoidal tracts,(4,18,20) portal/sinusoidal thrombosis,(21) acute hepatitis(20) and acute endothelitis.(19,20) In our series, the most common features identified include Kupffer cell hypertrophy in 21 (72.41%) patients,acute submassive hepatic necrosis (27.5%) followed by acute on chronic liver failure (13.7%) in a background of chronic liver diseases, features of NCPF (10.3%) and cholestasis (24%). Whether the changes of centrizonal hemorrhagic necrosis (13.7%) and macrovesicular steatosis (17.2%)were related to terminal congestive cardiac failure or patients’ metabolic syndrome, or were also contributed by virus-induced pathology could not be fully ascertained at this stage. Two of the cases of NCPF described above showed portal vein and sinusoidal thrombi (Figures 6D, K & L). However, the paucity of thrombosis in the rest of the cases of hepato-biliary system corroborated well with findings of studies done previously, as exemplified in a recent systematic review.(13)Though in our cohort of liver biopsies, hepatic lobular inflammatory cell infiltrate was not prominent, changes of hepatocyte degeneration as ballooning, acidophil bodies, MDBs(Mallory-Denk bodies), and microvesicular steatosis were prominent. Lack of inflammatory cell infiltrates in the liver has been described in earlier reports on fatal Covid-19 infections.(18) Our findings disagree with the observation of Sonzogni A et al.(21) that liver is not a primary target of Covid-19 infection and only vascular changes in liver are observed. In this cohort, we identified substantial histological changes of hepatocyte necrosis, degeneration, Kupffer cell hypertrophy, micro, and macrovesicular steatosis, more than the vascular changes.(21) The relative paucity of inflammation in the liver may be related to the treatment regimen or the inherited limitations of sampling by biopsies. Our histological findings match with those described by Lagana SM et al. except the fibrin ring granulomas the authors identified in 3 of their cases out of 40 liver biopsies examined(22). Out of the 8 liver biopsies which showed acute hepatic necrosis in our series, macrovescular steatosis was identified in 3 (37.5%) and microvescular steatosis was noted in another 3 biopsies (37.5%) [Supplementary Table 1]. A report from China on 300 fatal Covid-19 cases, it was found that steatosis and high neutrophil to lymphocyte ratio were the indicators of clinical aggressivenes.(23) This needs further elucidation. Except for paucity of the bile duct in one of our cases, the bile duct pathologies were not prominent in our cases, supporting the observation of Tian S. et al(4).

Renal histopathology in COVID-19 is reported to show changes including prominently acute tubular injury (more prominent in the proximal tubules)(18,24–28), arteriosclerosis (18) or glomerulosclerosis (29) (both as features of underlying comorbid conditions like hypertension), focal segmental glomerulosclerosis with a collapsing phenotype (25–27) and tubulointerstitial inflammation(24,26). Renal vascular changes reported are relatively less common and include fibrin thrombi (27,30), thrombotic angiopathy(29), and lymphocytic endothelitis (19). In our cohort of patients, the most common finding on renal histopathology was acute tubular injury and pre-existing renal conditions were only evident in 18%. None of the cases showed significant vascular changes.

Cardiac pathological examination in COVID-19 patients most commonly is reported to reflect underlying conditions like myocardial hypertrophy, interstitial fibrosis, and atherosclerosis.(4,18,19,28,31,32) Other findings described included atypical interstitial fibrosis,(4)myocardial edema,(4,19) and lymphocytic myocarditis.(33)(34) In our series only one patient had features of focal myocarditis.

This study highlighted the importance of MITS during a raging pandemic and demonstrated significant histopathological changes in fatal cases of Covid-19. The relative paucity of inflammatory cell infiltrates or fibrin thrombi in organs like liver, kidney, and myocardium may have been related to sampling limitations or therapy-related changes(anticoagulants). It was difficult to ascertain whether the pathological changes were entirely related to the viral pathology or reflected the comorbidities in the deceased patients. However, this study adds to the growing evidence that though pulmonary pathologies are most prominent in COVID-19 infections, extra-pulmonary organs like the liver and kidney are also affected, often remarkably.

## Data Availability

all data presented in the manuscript are available offline and can be referred to if necessary.

## Supplementary

**Table 1:** Clinical and histopathological diagnosis (n=37)

| SN | Age | Sex | Comorbidities | Ante-mortem diagnosis | Histopathological diagnosis |
| --- | --- | --- | --- | --- | --- |
| 1 | 40s | Female | Pulmonary and pleural tuberculosis (on treatment) | Acute Respiratory distress syndrome | Lung: DAD (exudative phase), bilateral non-small cell carcinoma |
| 2 | 60s | Female | Hypertension, Diabetes, Chronic kidney disease not on Dialysis | Acute Respiratory distress syndrome, acute kidney injury with existing chronic kidney disease | Lung: DAD (organizing phase), acute bronchopneumonia, megakaryocytes present , Heart: Marked hypertrophy, Liver : Centrizonal hepatocyte dropout with microvesicular steatosis., Kidney: mild focal Acute Tubular Injury with Diabetic Glomerulosclerosis |
| 3 | 80s | Male | Hypertension, Hypothyroidism | Acute Respiratory distress syndrome, acute kidney injury, encephalopathy, atrial fibrillation, ventricular tachycardia | Lung: DAD (organizing phase), acute bronchopneumonia, Heart : Hypertrophy |
| 4 | 30s | Female | Chronic myeloid leukaemia with blast crisis | Acute Respiratory distress Syndrome, hyperleukocytosis | Lung: DAD (exudative phase) |
| 5 | 20s | Male | Acute myeloid leukaemia with relapse | Acute Respiratory distress syndrome , acute kidney injury, lower respiratory tract infection with sepsis and septic shock | Lung: DAD (exudative phase), microthrombi present |
| 6 | 30s | Female | None | Acute Respiratory distress syndrome | Lung: DAD (exudative phase), Liver: Microvesicular steatosis&kuppfer cell hypertrophy |
| 7 | 50s | Male | Past Pulmonary tuberculosis | Acute Respiratory distress syndrome, | Lung: DAD (organising phase), acute |
|  |  |  |  | acute kidney injury, lower respiratory tract infection with sepsis and septic shock | bronchopneumonia, Heart: hypertrophy, Liver: Sub-massive centrilobular confluent necrosis with macrovesicular steatosis |
| 8 | 10s | Male | Aplastic Anaemia | Acute Respiratory distress syndrome with thrombocytopenia related bleed | heart: normal, Liver: Sub-massive centrilobular confluent necrosis |
| 9 | 60s | Male | Hypertension, Diabetes, Chronic liver disease (Child's class C) | Acute Respiratory distress syndrome, acute on chronic liver failure and hepatic encephalopathy, upper gastrointestinal bleeding | Lung: DAD (exudative phase), acute bronchopneumonia, Heart: Normal |
| 10 | 40s | Male | Chronic kidney disease on maintenance haemodialysis, Post Renal Transplant with graft rejection | Acute respiratory distress syndrome, intracranial haemorrhage, lower respiratory tract infection with sepsis, acute kidney injury with existing chronic kidney disease | Lung: Acute bronchopneumonia, Heart: normal, Liver: Sinusoidal and portal vein fibrin thrombi, Kidney: pre-existing pathology of Chronic rejection |
| 11 | 50s | Female | Hypothyroidism, diffuse cutaneous systemic sclerosis with interstitial lung disease | Acute respiratory distress syndrome | Lung: Acute lung injury, Heart: Normal, Liver: Diffuse macrovesicular steatosis |
| 12 | 60s | Female | Hypertension, Hypothyroidism | Acute respiratory distress with Myxoedema coma | Liver: Zone 3 & 2 macrovesicular steatosis |
| 13 | 20s | Male | Chronic kidney disease | Acute respiratory distress syndrome, Cytokine release syndrome, ventilator associated pneumonia, sepsis, acute kidney injury with existing chronic kidney disease | Lung: DAD (organising phase), acute bronchopneumonia, bacterial abscesses. microthrombi present. Heart: Focal myocardial calcification. Liver: Microvesicular steatosis. Kupffer cell hypertrophy. Kidney: Diffuse acute tubular injury. |
| 14 | 50s | Male | Hypertension, Diabetes, Coronary artery disease (old myocardial infarction) | Acute respiratory distress syndrome, ST elevation anterior wall myocardial infarction, cardiogenic shock, refractory ventricular tachycardia | Lung: DAD (exudative phase) with hyaline membrane |
| 15 | 10s | Male | Diffuse large B cell lymphoma | Acute respiratory distress syndrome, lower respiratory tract infection with sepsis and bacteraemia, CNS aspergillosis | Lung: DAD (organising phase) with microthrombi, Liver: Kupffer cell hypertrophy. Rest within normal limits. |
| 16 | 50s | Female | Hypothyroidism, Carcinoma endometrium, Chronic liver disease (non-alcoholic fatty liver disease) | Acute Respiratory distress syndrome, acute on chronic liver failure and hepatic encephalopathy, upper gastrointestinal bleeding | Lung: DAD (organising phase) with microthrombi, Liver: Architectural distortion, mixed inflammation in portal tracts with focal interface hepatitis, nodular centrilobular necrosis with macrovesicular steatosis, numerous acidophil bodies, Kupffer cell hypertrophy, portal vein showing a recanalized thrombus |
| 17 | 60s | Female | Carcinoma lung | Carcinoma lung with lower respiratory tract infection (post-obstruction), with sepsis and septic shock, acute kidney injury | Lung: primary adenocarcinoma (TTF-1 negative). DAD with hyaline membrane. Heart: normal. Liver: Centrilobular necrosis. Kupffer cell hypertrophy. |
| 18 | 20s | Male | Hypertension, Chronic kidney disease, Abdominal aortic aneurysm | Acute respiratory distress syndrome, Cytokine release syndrome, ventilator associated pneumonia, sepsis, acute kidney injury with existing chronic kidney disease | Lung: Acute lung injury with alveolar hemorrhage. Liver- Centrilobular hepatocyte necrosis with ballooning and diffuse microvesicular steatosis. Kupffer cell hypertrophy. |
| 19 | 60s | Female | Carcinoma tongue | Acute respiratory distress syndrome, haemorrhagic shock due to warfarin induced coagulopathy | Lung: DAD (exudative phase) Liver: Kupffer cell hypertrophy, focal interface hepatitis |
| 20 | 40s | Male | Hypertension, Diabetes, Chronic kidney disease on maintenance haemodialysis, Post-Cardio-pulmonary-resuscitation | lower respiratory tract infection with sepsis, post-cardiopulmonary resuscitation | Lung: Focal fibrin deposit<br>Liver: Mild lymphocytic infiltrate with focal absence of bile ducts, cholestasis, Kupffer cell hypertrophy, and focal perisinusoidal fibrosis.<br>?idiopathic non-cirrhotic portal hypertension |
| 21 | 50s | Female | Rheumatic Heart Disease, severe Pulmonary artery hypertension, atrial fibrillation with fast ventricular rate | Acute respiratory distress syndromes, acute kidney injury, refractory ventricular arrhythmias | Lung: DAD (organizing phase), acute bronchopneumonia, Liver: Zone 3 necrosis with macrovesicular steatosis, Kidney: Diffuse acute tubular injury and vacuolization. Mild arterial fibro-intimal expansion |
| 22 | 30s | Male | Hypertension, Diabetes, Hypothyroidism, Chronic kidney disease not on Dialysis, Post renal transplant with chronic graft rejection, Cytomegalovirus infection (kidney) | Acute respiratory distress syndrome, lower respiratory tract infection, sepsis with septic shock, acute kidney injury on chronic kidney disease | Lung: N/A, Liver: Diffuse ballooning with focal necrosis, acidophil bodies, and focal peri-sinusoidal space and central vein thickening. ?idiopathic non-cirrhotic portal hypertension. Kidney: Diffuse acute tubular injury, 50% IFTA with focal microcalcifications, |
| 23 | 40s | Male | Hypertension, Chronic kidney disease on maintenance haemodialysis | Acute respiratory distress syndrome, basal ganglia haemorrhagic infarct with midline shift and raised intracranial pressure | Lung: DAD (exudative phase), acute bronchopneumonia, Liver: Diffuse interface hepatitis with focal confluent necrosis, Kupffer hypertrophy, secondary hemosiderosis. |
| 24 | 60s | Male | None | Acute respiratory distress syndrome, cytokine release syndrome, lower respiratory tract infection, sepsis with septic shock | Lung: Acute lung injury, acute bronchopneumonia, Type II pneumocyte hyperplasia with atypia, Liver: Mild lymphocytic infiltrate with patchy ballooning and necrosis, centrilobular macrovesicular steatosis, focal canalicular cholestasis, Kupffer cell hypertrophy. Kidney: Acute tubular injury, white blood cell casts, Arterial intimal fibrosis, <5% IFTA |
| 25 | 60s | Male | Hypertension, Diabetes | Acute respiratory distress syndrome, Cytokine release syndrome, lower respiratory tract infection, sepsis with septic shock, acute kidney injury | Kidney: Diffuse acute tubular injury, extensive arteriolar hyalinosis. |
| 26 | 20s | Female | Chronic liver disease, Autoimmune hepatitis | Acute respiratory distress syndrome, acute on chronic liver failure, upper gastrointestinal bleeding, hepatic encephalopathy, spontaneous bacterial peritonitis, sepsis with septic shock | Lung: DAD (exudative phase), acute bronchopneumonia, Liver: Distorted architecture, Acute on chronic liver failure, sub-massive hepatic necrosis, Features suggestive of autoimmune hepatitis, and autoimmune cholangitis (overlap). |
| 27 | 50s | Male | Alcohol user, Chronic liver disease | Acute respiratory distress syndrome, coagulopathy, upper gastrointestinal bleeding, ventilator associated pneumonia, sepsis with septic shock, subdural haematoma, acute kidney injury | Lung: Acute organising pneumonia, pulmonary oedema, Liver: Focal mild interface hepatitis, macrovesicular steatosis, Kupffer cell hypertrophy, diffuse ballooning, foci of spotty necrosis, extensive MD bodies, and necrotic hepatocytes., Kidney: Diffuse acute tubular injury. |
| 28 | 60s | Female | Aplastic anaemia, Past Pulmonary tuberculosis | Acute respiratory distress syndrome, cytokine release syndrome, lower respiratory tract infection, sepsis with septic shock, acute ischemic stroke | Lung: Acute organising pneumonia, alveolar haemorrhage, microthrombi, megakaryocyte, hyperplastic pneumocytes, Liver: Mild chronic hepatitis with secondary hemosiderosis and irregular macrovesicular steatosis Kidney- diffuse acute tubular injury. |
| 29 | 30s | Male | Diabetes | Pulmonary tuberculosis (active), pulmonary mucormycosis, unilateral secondary spontaneous pneumothorax | Lung: Acute lung injury, acute bronchopneumonia, Liver: Irregular macrovesicular steatosis, Kidney: diffuse acute tubular injury with focal interstitial nephritis. |
| 30 | 80s | Female | Diabetes, Coronary artery disease, Dilated cardiomyopathy, atrial fibrillation with | Gastroenteritis (?COVID related) with sepsis with septic shock, acute kidney | Lung: normal lung tissue, Liver: sub-massive hepatic necrosis with microvesicular steatosis. |
|  |  |  | controlledventricular rate | injury | PT: no thrombi/obliteration, |
| 31 | 50s | Male | Hypertension, Diabetes, Chronic kidney disease on maintenance haemodialysis | Acute respiratory distress syndrome, Cytokine release syndrome, lower respiratory tract infection, sepsis, acute kidney injury with existing chronic kidney disease, right pneumothorax | Lung: Acute Organising Pneumonia, Acute Bronchopneumonia with microthrombi, Liver: Foci or irregular hepatocyte necrosis |
| 32 | 60s | Male | Hypertension, Diabetes | Acute respiratory distress syndrome, Cytokine release syndrome, lower respiratory tract infection, sepsis with septic shock, acute kidney injury | Liver: Zone 2 and 3 hepatocyte necrosis with dense polymorph infiltrate, Zone 1 focal hepatocyte necrosis, centrilobular macrovesicular steatosis (25 %) with nuclear glycogenization-sub-massive hepatic necrosis |
| 33 | 60s | Male | Alcohol user, Chronic liver disease, Hilar cholangiocarcinoma | Acute respiratory distress syndrome, cholangitis with sepsis with septic shock with septic encephalopathy, acute kidney injury | Lung: Normal lung with interalveolar capillary dilation<br>Liver: Moderate mixed inflammation in portal tracts and focal interface hepatitis; ascending cholangitis and features of sepsis; Hepatocytes-extensive intracanalicular cholestasis, focal ballooning, and an occasional hepatocyte dropout at Z3. |
| 34 | 60s | Male | Hypertension, Diabetes, Chronic kidney disease not on Dialysis, Thalamic infarct | Acute respiratory distress syndrome, acute kidney injury with existing chronic kidney disease, coagulopathy, raised intracranial pressure | Lung: DAD (organising phase),acute bronchopneumonia, microthrombi.<br>Liver- mostly normal |
| 35 | 10s | Male | T-cell acute lymphoblastic leukaemia (on treatment) | Acute respiratory distress syndrome, Cytokine release syndrome, Gastroenteritis (?COVID related) with neutropenic sepsis with septic shock, ?septic encephalopathy | Lung: Engorged capillaries |
| 36 | 30s | Female | Hypertension, Hypothyroidism, Chronic kidney disease on maintenance haemodialysis, Chronic liver disease | Acute respiratory distress syndrome, lower respiratory tract infection, sepsis with septic shock, hepatic encephalopathy, acute kidney injury with existing chronic kidney disease, coagulopathy | Lung: chronic interstitial pneumonia<br>Liver: Architecture: distorted. Dense mixed inflammation in portal tracts with focal interface hepatitis. Marked bile ductular proliferation, features of sepsis, ductular cholestasis; Hepatic nodules extensive confluent necrosis within the hepatic nodules, cholestasis, microvesicular steatosis and Spotty necrosis. |
| 37 | 30s | Female | Hypothyroidism | Lower respiratory tract infection, Acute bacterial meningitis, sepsis with septic shock, acute kidney injury | Lung: Chronic interstitial inflammation<br>Liver: Mild chronic inflammation in portal tracts with focal interface hepatitis. Hepatocytes normal. |

